# Nonmyeloablative Pentostatin-Cyclophosphamide Preconditioning Improves Rates of Engraftment in Adults Undergoing Haploidentical HCT for Sickle Cell Disease

**DOI:** 10.1101/2025.08.31.25334807

**Authors:** Emily Limerick, Matt Hsieh, Jackie Queen, Mary Lacy Grecco, Julia Varga, Jennifer Brooks, Wynona Coles, Neal Jeffries, Courtney Fitzhugh

## Abstract

The morbidity and mortality of sickle cell disease (SCD) remain high. Novel nonmyeloablative haploidentical hematopoietic cell transplant (HCT) regimens are being proposed.

**Methods:** This report compared the effect of adding an oral cyclophosphamide and IV pentostatin (PC) preconditioning to the nonmyeloablative NIH HCT platform of alemtuzumab and total body irradiation (TBI).

**Results:** Thirty-nine adult SCD patients were included. The median age was 33 years, 61% were male, and 92% were HbSS genotype. The median follow-up was 6.5 years. Many patients had severe end-organ damage, including dialysis-dependent end-stage kidney disease (10%) and cirrhosis (8%).

One-year overall survival was 95%. The PC regimen was associated with a reduction in acute rejection one-year post-HCT (5% vs. 44%; p=0.004) and lower graft failure rates throughout the follow-up period. After a median follow-up of 5.2 years, the disease-free survival was 71% for the PC regimen. The PC preconditioning was associated with higher rates of full donor chimerism at 2-years post-HCT (0% vs. 43%; p=0.02). Grade II-IV acute graft-versus-host disease (GVHD) rates were low; no patients developed moderate to severe chronic GVHD. There remain no cases of myeloid malignancy after PC. With the increased immunosuppression of PC, 23% of patients developed post-transplant lymphoproliferative disorder, 19% developed immune cytopenias, and 62% had viral reactivation.

**Summary:** Further study to determine an optimal nonmyeloablative haploidentical regimen for SCD patients with compromised organ function is imperative.

## Introduction

Sickle cell disease (SCD) is associated with considerable morbidity and mortality. We and others have shown that adults with organ dysfunction are more likely to die at least 20 years earlier than the general population [1-10]. And those with more than one organ involved are expected to die even earlier[11]. Despite decreased mortality for children with SCD [12-14], the median age of death remains within the 5^th^ decade of life[3].

Hematopoietic cell transplantation (HCT) offers a curative option for individuals with SCD. Myeloablative HLA-matched sibling donor (MSD) HCT is efficacious [15, 16] but this approach is limited to younger patients with minimal organ damage. Nonmyeloablative MSD HCT allows the enrollment of patients with compromised organ function. While the NIH approach has minimal GVHD and transplant-related mortality, the disease-free survival (DFS) has been around 85-88% [17, 18]. Further, <15% of patients with SCD have HLA-MSD [19].

Autologous gene therapy and gene editing strategies are being developed; the Food and Drug Administration recently approved two approaches [20, 21]. While both strategies showed remarkable efficacy, currently, there is a requirement for myeloablative chemotherapy, substantially limiting access for many adults with SCD given the increased prevalence of hepatopathy, nephropathy, and cardiopulmonary disease with age.

At least 90% of patients with SCD have a parent, child, half-matched sibling, or other relative who could serve as a haploidentical donor [22]. Thus, nonmyeloablative haploidentical HCT could be extended to most adults with SCD, including those with compromised organ function. Our first nonmyeloablative haploidentical HCT study was written as a dose escalation of post-transplant cyclophosphamide (PT-Cy) [23]. All patients received alemtuzumab, 400cGy total body irradiation (TBI), and sirolimus. Adding PT-Cy improved the engraftment rate and DFS with no significant GVHD. However, the DFS was, at best, 50%. In addition, the incidence of myeloid malignancies was unacceptably high, with 3 of 21 patients experiencing myeloid malignancies. The study closed to accrual in 2016.

We and others have worked to optimize nonmyeloablative haploidentical HCT to widen the donor pool with acceptable rates of chronic GVHD and mortality [16, 24-30]. Traditionally, our goal has been mixed donor/recipient chimerism to minimize the risk of GVHD, as we found that 20-25% donor myeloid chimerism (DMC) is sufficient to reverse the sickle phenotype [31, 32]. Others with a therapeutic goal of full donor chimerism have had excellent DFS and low GVHD rates [16, 27] but often excluded patients with severe organ damage and had limited long-term follow-up.

Because of the high rate of acute graft rejection with our original haploidentical HCT study[23], we hypothesized that additional immunosuppression targeting recipient lymphocytes was necessary. Others have shown that pentostatin combined with Cy (PC) results in significantly higher recipient T- and B-cell depletion than fludarabine and Cy (FC) in a murine model [33]. Importantly, mice receiving PC engrafted at a higher rate and more commonly displayed stable mixed chimerism than FC. In a pilot HCT study in 10 patients with metastatic renal cell carcinoma using PC conditioning [34], recipient T-cells were effectively depleted without severe neutropenia; all 10 patients engrafted with stable mixed chimerism, even after discontinuing sirolimus. We, therefore, sought to develop a nonmyeloablative haploidentical HCT regimen that included PC preconditioning (haplo-PC), which would maximize long-term efficacy while minimizing toxicity, and could be offered to patients with compromised organ function. We also strove to evaluate whether the addition of PC conditioning impacted transplant outcomes, including the acute graft rejection rate, overall survival, DFS, the incidence of GVHD and viral complications, and the timing of lymphocyte reconstitution, compared to the original haploidentical protocol.

### Materials and Methods

This report compares the results of NIH’s two nonmyeloablative haploidentical HCT regimens. The National Heart, Lung, and Blood Institute Institutional Review Board approved both studies. All patients gave written informed consent. An independent data and safety monitoring board monitored the studies. Our first haploidentical protocol (haplo-1; NCT00977691) employed alemtuzumab and TBI-based conditioning with dose-escalating PT-Cy and sirolimus for GVHD prophylaxis in adults with severe SCD; the results of this study have been previously reported [23]. While all patients are included in descriptive reporting of significant adverse events, these analyses excluded patients who did not receive PT-Cy (n=3); patients who received repeat transplants were censored at the time of repeat HCT.

The second study (haplo-PC) was a prospective phase 1/2 nonmyeloablative haploidentical peripheral blood stem cell (PBSC) transplant protocol for patients with severe SCD (NCT03077542). The initial inclusion criteria included any type of SCD with history of stroke, tricuspid regurgitant velocity (TRV) >2.5 m/s with or without right heart catheterization (RHC)-documented pulmonary hypertension [7], sickle hepatopathy [35], defined as ferritin >1000 mcg/L and platelet count <250,000/µL or direct bilirubin >0.4 mg/dL and platelet count <250,000/µL; patients with frank cirrhosis, defined based on liver biopsy, were also included, recurrent vaso-occlusive crises, or recurrent acute chest syndrome. Due to a higher-than-expected rate of complications, the details of which will be discussed in this report, the inclusion criteria were updated in 2022 to only include patients with higher risk disease: stroke, TRV >2.7 m/s with or without pulmonary hypertension, sickle hepatopathy (as defined above), acute chest syndrome requiring intensive care admission, or silent infarcts; patients with recurrent vaso-occlusive crises in the absence of other organ manifestations were no longer included. The recruitment period for the study started on 2/7/2017 and ended on 6/23/2023.

At the time of enrollment, patients had to be at least 18 years of age, have a haploidentical related donor available, ejection fraction ≥ 35%, glomerular filtration rate (GFR) >60 mL/min/1.73 m^2^ (this was due to the addition of pentostatin and differs from the haplo-1 protocol in which there was no exclusion based on renal function; indeed patients with dialysis-dependent end-stage kidney disease (ESKD) were included on haplo-1) and adjusted diffusing capacity of the lungs for carbon monoxide >35%. Patients with donor-specific anti-HLA antibodies with a mean fluorescence intensity >2000 [36] and recipients seronegative for Epstein-Barr Virus (EBV) who had EBV seropositive donors [37] were excluded.

All patients received PC preconditioning with 4 doses of intravenous pentostatin (4mg/m^2^) given on days -21, -17, -13, and -9 (Figure 1). Estimated GFR (eGFR) was calculated before each dose of pentostatin, and the dose was adjusted as necessary. Although an eGFR at time of protocol enrollment >60 mL/min/1.73 m^2^ was required, pentostatin is associated with a risk of severe renal toxicity. Table S1 summarizes the pentostatin dose adjustment schema as necessary after pentostatin initiation. Oral Cy was given on day -21 until day -8 at 200mg/day. To maximize recipient lymphodepletion, the goal absolute lymphocyte count (ALC) before alemtuzumab was <100/µL. The goal absolute neutrophil count (ANC) before alemtuzumab was >1,000/µL to prevent excessive and prolonged myelosuppression. Cy dosing was adjusted per protocol to achieve the desired ALC and ANC goals (Table S2).

**Figure 1.**
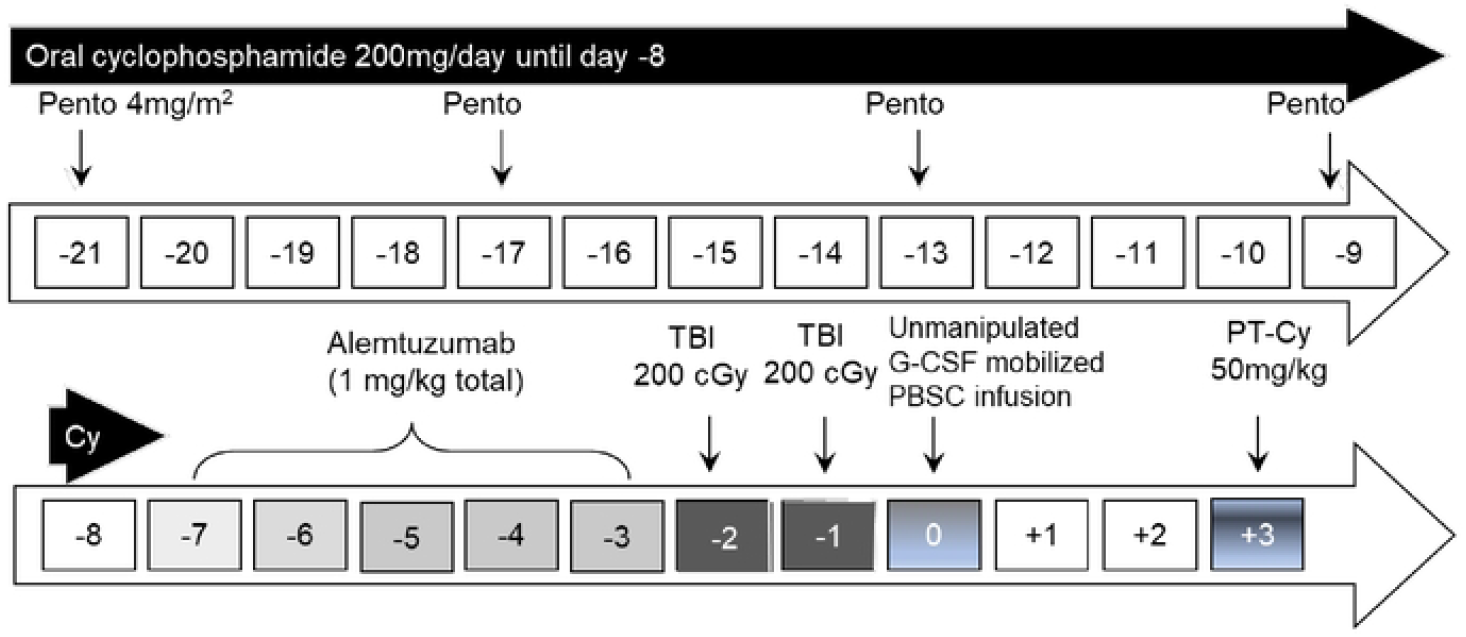
Conditioning regimen schema. Pento: pentostatin; TBI: total body irradiation; G-CSF: granulocyte colony-stimulating factor; PBSC: peripheral blood stem cell; PT-Cy: post-transplant cyclophosphamide.

Patients received a total 1mg/kg of alemtuzumab infused on days 7-3 before PBSC infusion. The first 20 patients received 400 cGy TBI in 2 divided doses on days –2 and –1 before PBSC infusion; patients 20 - 22 received 400 cGy TBI as a single fraction. Post-transplant cyclophosphamide was infused as a single dose of 50 mg/kg on day +3 based on our prior experience of higher rates of viral reactivation with two doses[23]. A sirolimus trough level of 10 to 12 ng/mL was targeted until 3 to 4 months post-HCT, and then the level was decreased to 5 to 10 ng/mL in engrafted patients. Sirolimus was generally continued as tolerated in engrafted patients until at least 1-year post-HCT or until the patients achieved full donor chimerism (>95% DMC and >95% donor lymphoid chimerism (DLC)).

Recipients positive for cytomegalovirus (CMV) received letermovir prophylaxis from day -21 through 100 days post-HCT. The goal hemoglobin level after the start of alemtuzumab was 9 g/dL and a platelet count of 50,000/µL. Additional supportive care was provided as previously described [23] and per the protocol. Donors received 5 to 6 days of granulocyte colony-stimulating factor at 10 to 16 μg/kg/d for mobilization, followed by peripheral blood leukapheresis with a goal of at least 10 × 10^6^ CD34 cells per kilogram of the recipient’s body weight. The PBSC products were subsequently cryopreserved [38].

We examined rates of graft rejection at one year post-HCT and graft failure at the last follow-up. Graft rejection refers to an acute loss of the graft, with <5% DMC and DLC. Graft failure includes those with graft rejection but also includes patients with return of acute complications of SCD, regardless of whether donor cells remained detectable. Planned follow-up occurred 1 to 2 times per week until 100 days post-HCT, every 6 months until 2 years post-HCT, and annually thereafter.

Laboratory studies, including hematologic studies, chemistries, and lymphocyte subsets, were run at the NIH Department of Laboratory Medicine. Lineage-specific chimerism studies (CD14/CD15 and CD3) were conducted as previously described [39, 40]. Donor red cell chimerism was assessed by hemoglobin electrophoresis and extended red blood cell (RBC) phenotyping; the NIH Department of Transfusion Medicine performed RBC phenotyping and HLA antibody analyses.

The cumulative incidence of graft rejection at one-year post-HCT and graft failure at last follow-up were computed for each protocol, treating death without a prior graft failure as a competing risk. Tests of the equivalence of cumulative incidence between the two protocols were based on Gray’s test [41] and implemented using the cmprsk package in the R statistical language. Kaplan-Meier analyses of DFS do not require accounting for competing risk, as both death and graft failure count as events—this analysis was implemented in the survival package of the R language. Comparisons of categorical outcomes between protocols were made with Fisher’s exact test, and continuous variables with the Wilcoxon rank sum test.

## Results

The cohort included 39 patients who were predominantly male (61%) and HbSS genotype (92%) (Table 1). The mean recipient age on the haplo-1 protocol was 36 years, while the mean recipient age on the haplo-PC protocol was 32 years. The mean donor age was 45 and 33 years on haplo-1 and haplo-PC, respectively. There were 18 (46%) patients on haplo-1 and 21 (54%) on haplo-PC. Most patients (90%) had a history of hydroxyurea use, and a substantial portion (38%) required regular transfusion therapy prior to HCT. Clinical complications were numerous, including elevated TRV (62%), RBC alloimmunization (46%), acute chest syndrome requiring ICU admission (41%), sickle hepatopathy (38%), and stroke (36%) (Table 1). This cohort included patients often excluded from other curative therapies, including those with cirrhosis (n=3) and ESKD requiring dialysis (n=4). Indeed, 37% of US patients in this cohort were ineligible for what is becoming the most common haploidentical HCT approach for adults with SCD [27, 42, 43]. There was a significantly higher proportion of patients with sickle nephropathy on haplo-1 than haplo-PC; otherwise, baseline donor and recipient characteristics were similar between the two protocols.

One-year overall survival was 95%; in the first year, there was 1 death on each protocol. Table 2 summarizes the post-HCT outcomes. At one-year post-HCT, there was more acute rejection on the haplo-1 study compared to the haplo-PC group (44% vs. 5%, p=0.004) (Figure 2). The median follow-up was longer for the haplo-1 than the haplo-PC cohort (10.1 vs. 5.2 years; p<0.0001). The improved rates of sustained engraftment in the haplo-PC cohort were maintained throughout the follow-up period. We compared graft status at last follow-up: for patients who died during the follow-up period, we report their graft status at the time of death. Figure 3 shows 86% of patients had sustained engraftment at last follow-up in the haplo-PC cohort compared with 33% in haplo-1 (p=0.006). Two haplo-PC patients with slowly falling chimerism are being closely followed. Their most recent DMC levels were 15 and 16%, while hemoglobin electrophoresis showed 49.5 and 39.7% HbS at 5 and 7 years post-HCT, respectively; both donors have sickle cell trait. Overall survival at last follow-up was 67% on haplo-1 and 86% on haplo-PC. Figure 4 demonstrates that the DFS at the last follow-up was 33% (haplo-1) and 71% (haplo-PC); these differences show a trend towards improvement in DFS with the addition of PC (p=0.06). Five patients on the haplo-1 trial had repeat transplants in the setting of return of SCD: one died of intracranial hemorrhage 60 days after her repeat HCT in the setting of thrombocytopenia while receiving rituximab for EBV-associated post-transplant lymphoproliferative disorder (PTLD); another died from COVID-19 less than 1 year from the second transplant; a third patient who experienced graft failure received the transplant for AML and died of relapse. The remaining two repeat HCT patients are alive and free of SCD. Only one patient on the haplo-PC trial received a repeat transplant; that patient is alive and free of SCD.

**Figure 2.**
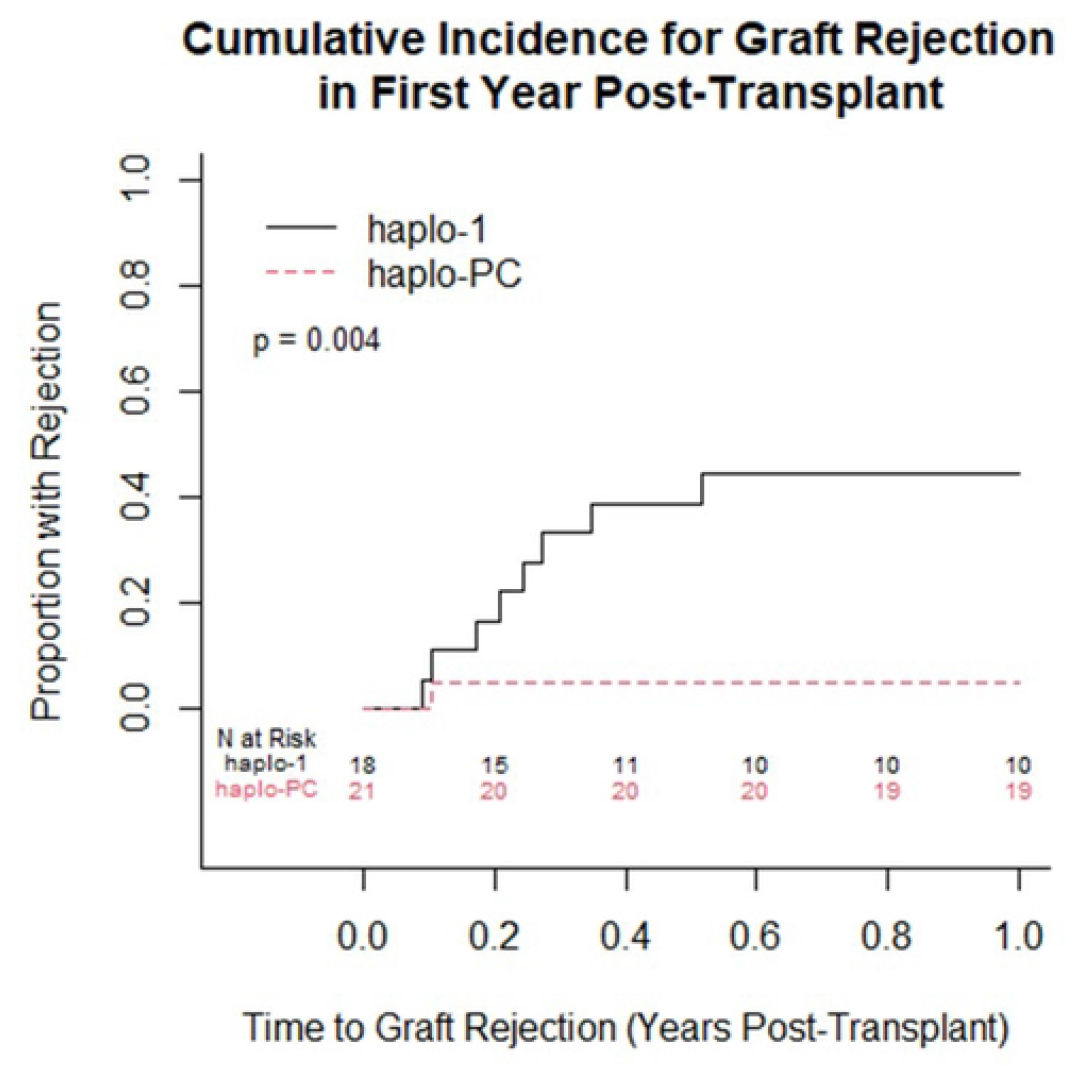
Cumulative incidence curves comparing the proportion with acute graft rejection within one year post-transplant between the 2 protocols (p=0.004).

**Figure 3.**
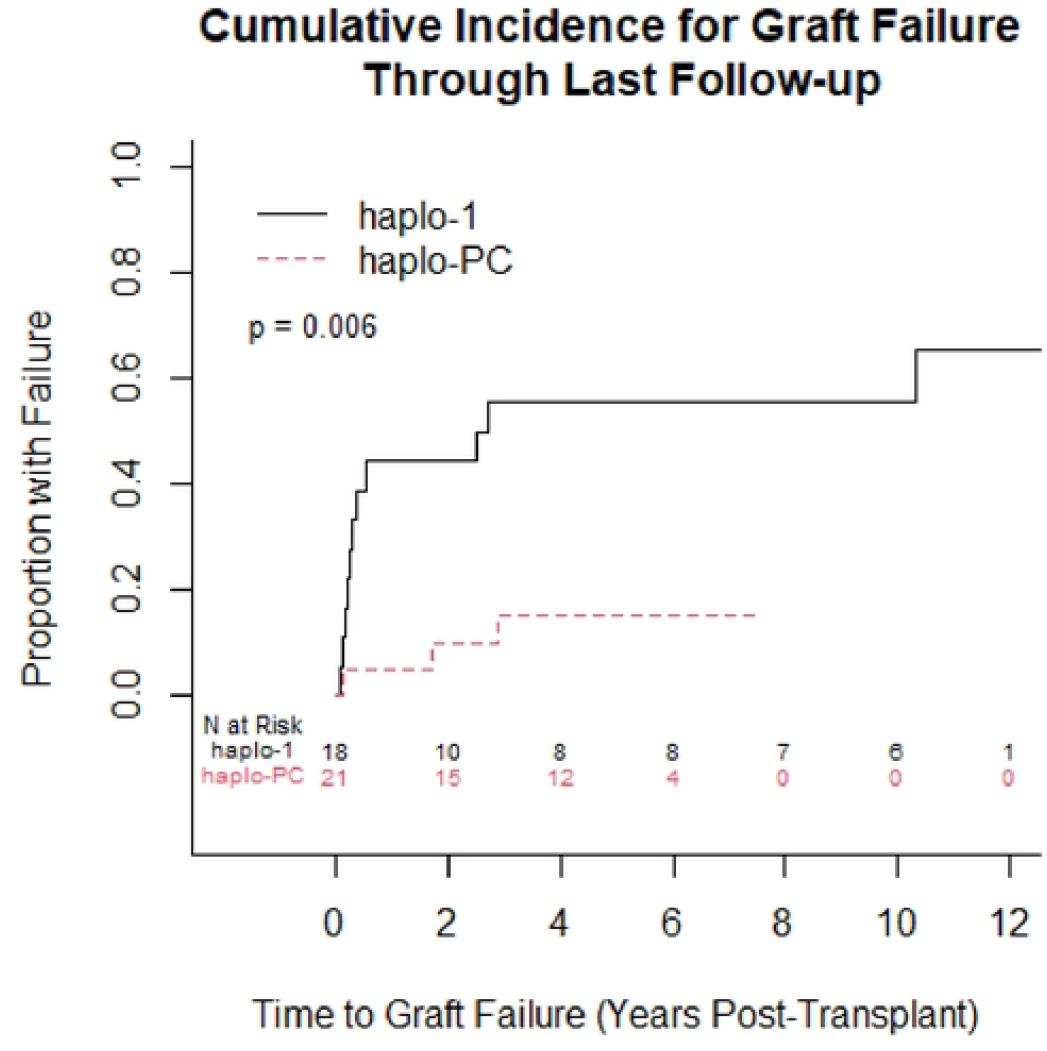
Cumulative incidence curves comparing the proportion with graft failure at last follow-up between the 2 protocols (p=0.006).

**Figure 4.**
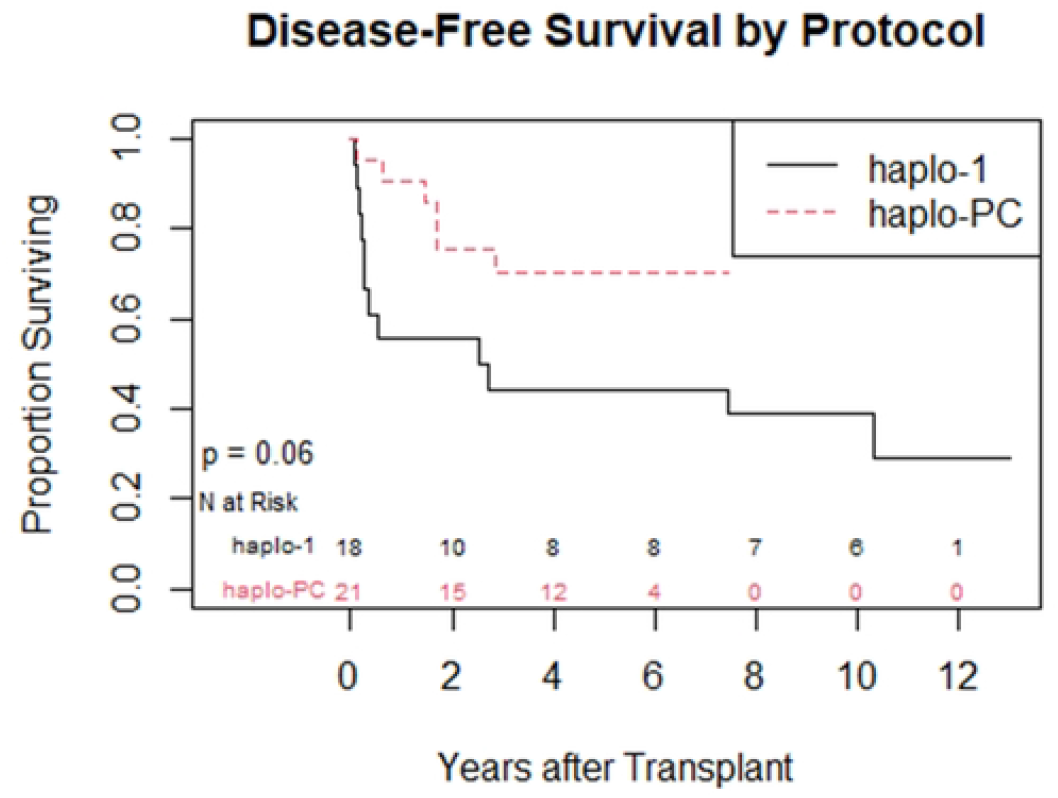
Kaplan Meyer curve comparing sickle cell disease-free survival at last follow-up between the 2 protocols.

No patients on the haplo-1 protocol and two (10%) patients on the haplo-PC protocol developed grade II-IV acute GVHD (one grade II, one grade IV). No patients on either protocol developed moderate to severe chronic GVHD.

We examined rates of full donor chimerism compared to mixed chimerism in patients free of SCD at 2-years post-HCT. On haplo-1, 0 of 10 (0%) patients had full donor chimerism while 6 of 14 (43%) haplo-PC patients had full donor chimerism (p=0.02). Figures 5a and 5b compare the mean donor chimerism levels between the protocols.

**Figure 5.**
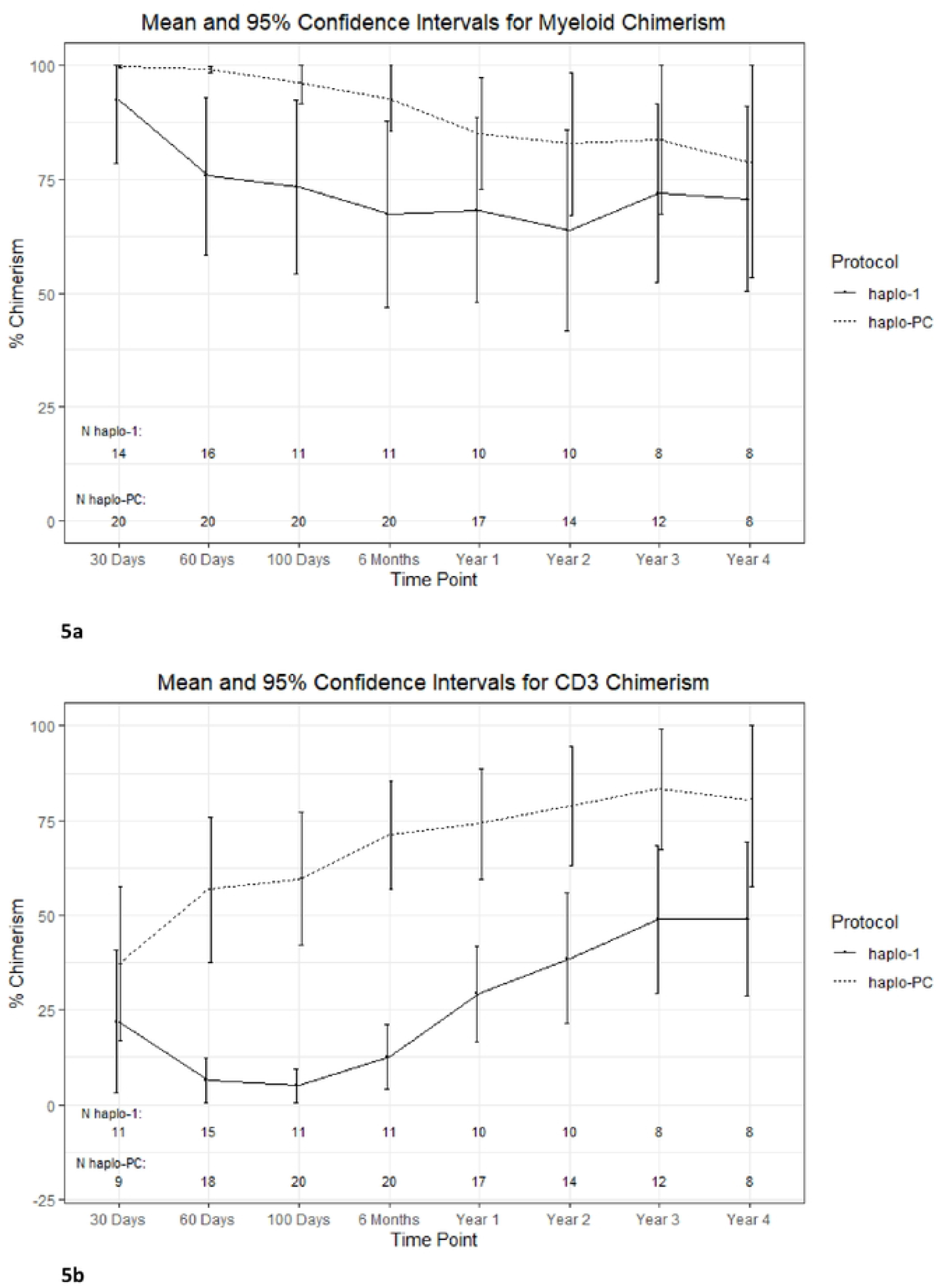
Percent donor chimerism trends 4 years after transplant a) Myeloid chimerism and b) CD3 chimerism

Of the patients who were alive and free of SCD at last follow-up, 4 (67%) haplo-1 patients and 11 (73%) in the haplo-PC cohort were off immunosuppression. Table 2 summarizes complications and outcomes after HCT. While 16 (89%) of haplo-1 patients experienced an episode of febrile neutropenia, ten (48%) patients on haplo-PC had a neutropenic fever early post-HCT (p=0.008). One patient on the haplo-1 protocol and two patients on haplo-PC experienced sepsis early after HCT (6% and 10% for haplo-1 and haplo-PC, respectively). There was a trend towards more viral reactivation (EBV, CMV) on the haplo-PC protocol than haplo-1: 62% versus 28% (p=0.05). Viral disease (adenovirus and CMV) rates were 17% and 0%, on haplo-1 and haplo-PC, respectively (p=0.21).

Three of 21 (14%) patients who received HCT on haplo-1 experienced myeloid malignancies, and no patients on haplo-PC (0%). However, as one patient from haplo-1 was excluded from the analysis since she did not receive PT-Cy, for the purpose of this analysis, 2 patients on haplo-1 developed a myeloid malignancy (11%) versus 0 patients on haplo-PC (p>0.05). Five of 22 patients (23%) who underwent HCT on haplo-PC developed PTLD, and no patients on haplo-1 (0%). Two patients developed severe, refractory PTLD treated with systemic chemotherapy and virus-specific T-cells. These therapies led to remission of PTLD in both patients, though one died of complications of therapy (methotrexate leukoencephalopathy). Three patients received rituximab alone, one of these patients, as previously noted, died secondary to intracranial hemorrhage. For the analysis, this one patient, who developed PTLD after a repeat HCT on haplo-PC (after her initial HCT on haplo-1), was excluded, and 4 patients on haplo-PC developed PTLD (19%) versus 0 patients on haplo-1 (p>0.05).

There were more immune complications, including Evans syndrome (n=2) and autoimmune hemolytic anemia (n=2; Table S3), in the haplo-PC protocol than haplo-1: 19% versus 0%, respectively (p=0.11). While these differences are not statistically significant, in part due to the small sample size, they are indeed clinically important. The presence of two lethal cases of Evans syndrome contributed to the modification of the protocol’s inclusion criteria and TBI administration in 2022. Although data suggest that DLC changes at 6 months may predict the subsequent development of immune cytopenias[44], we saw no such changes in our population (Figure S1).

Two patients on haplo-PC had biopsy-proven cirrhosis. Both tolerated the conditioning without significant event; neither experienced graft failure and both had full donor chimerism at last follow-up; one of these patients developed EBV-driven PTLD and died due to complications of PTLD therapy. Only one patient had detectable donor-specific HLA antibodies prior to HCT, though at the time of transplant they were no longer detectable. The patient remains engrafted and free of SCD at last follow-up. Eighteen (46%) patients had a reported history of alloimmunization prior to HCT; 17 (44%) had a specific RBC antibody identified at some point prior to HCT. There were three (8%) patients who received HCT from donors against whose RBC antigens they had a documented RBC antibody; none of these patients experienced graft failure.

The additional immunosuppression of the PC pre-conditioning in the haplo-PC protocol did not delay lymphocyte reconstitution. Figure 5 shows that the expected drop in lymphocyte counts in the early post-HCT period is similar between groups. Further, both protocols demonstrate a return to pre-HCT CD3, CD4, CD8, and CD19 counts by two years (Figure 6a-d).

**Figure 6.**
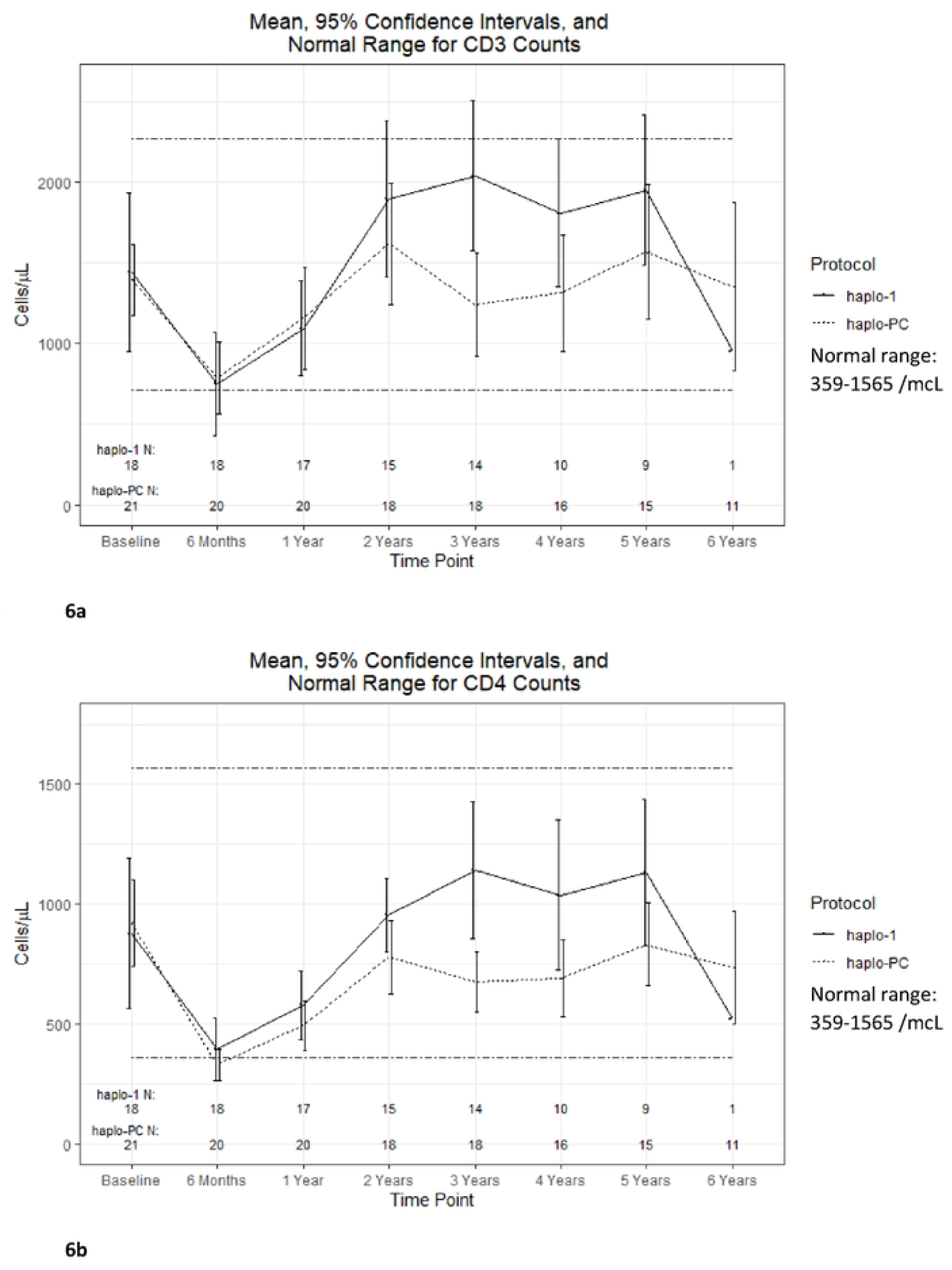
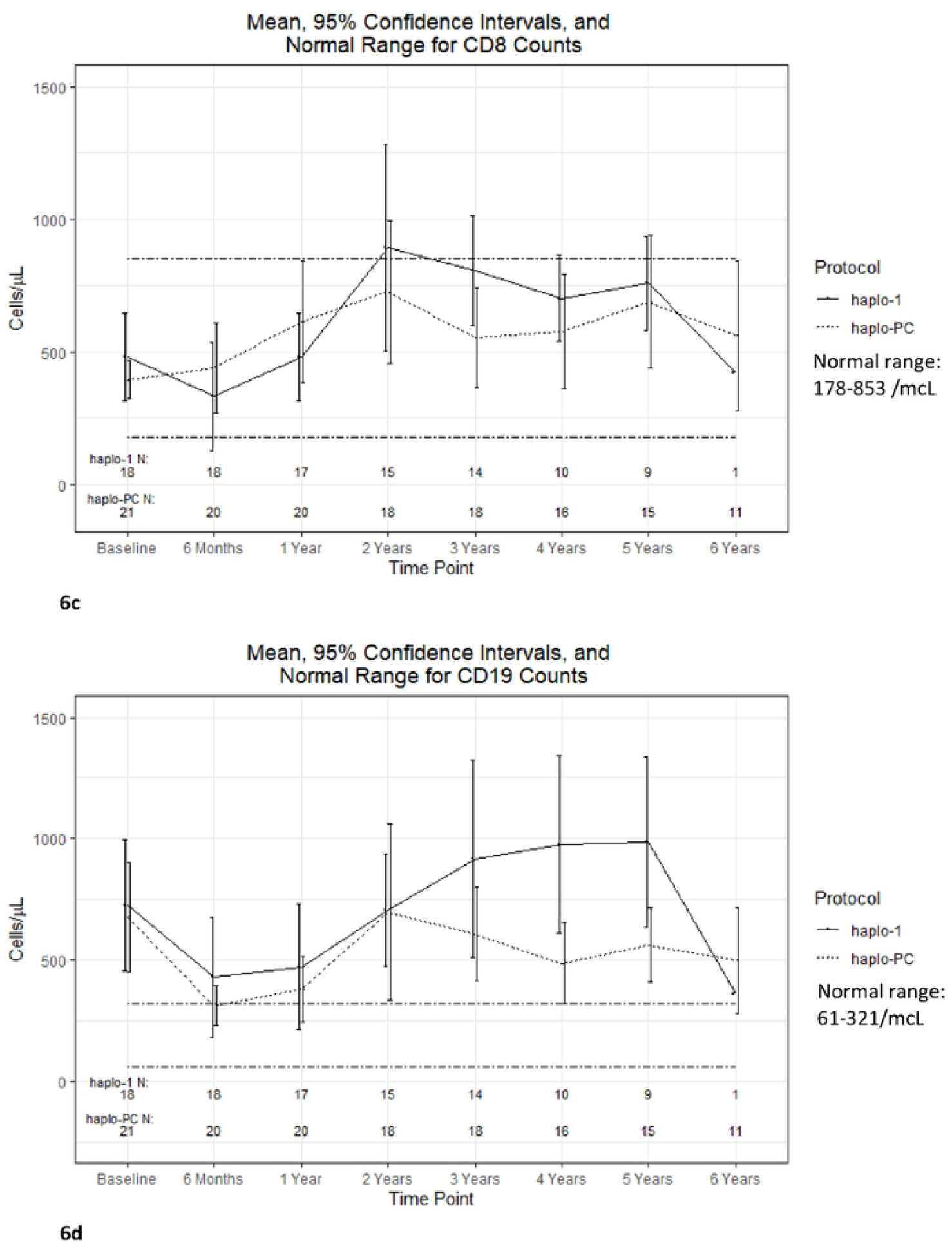
Comparison of mean lymphocyte reconstitution trends after HCT a)CD3 counts b)CD4 counts c)CD8 counts d)CD19 counts

## Discussion

Outcomes of haploidentical HCT for SCD have improved markedly since early studies, which were marred by high rates of graft rejection[22, 23]. Indeed, 88-95% event-free survival at 2 years for adults has been reported after bone marrow transplantation with reduced-intensity fludarabine, cyclophosphamide, ATG, thiotepa, and 200cGy TBI conditioning [27, 30, 45]. These excellent results rival the success of nonmyeloablative MSD HCT[17]. Murine studies demonstrate that the PC is nonmyeloablative, with significantly less marrow ablation than lethal doses of TBI despite profound immune depletion[33]. The regimen is nonmyeloablative, but consideration of toxicities remains important.

The impact of SCD comorbidities is dire: multi-system end-organ impairment was associated with a 4.2-fold increase in mortality risk[11]. Thus, access to potentially curative therapies that include patients with severe disease and end-organ damage is critical. The minimally toxic NIH regimens have been employed in patients often excluded from other protocols, including those with ESKD requiring dialysis, cirrhosis, and severe pulmonary hypertension[23, 46]. Indeed, 37% of US patients we transplanted would have been excluded from the Blood and Marrow Transplant Clinical Trials Network 1507 study, which employs a regimen identical to the Vanderbilt study. Thus, there remains an urgent need for efficacious HCT regimens for adults with SCD and severe end-organ damage.

As expected based on the murine and non-SCD clinical data described above [32, 42], as well as recently published data in the matched sibling HCT for SCD setting [47], we demonstrate that PC substantially reduced the high rates of acute graft rejection, likely due to the intensified lymphodepletion and host t-cell suppression. Late graft failures presented a challenge on both protocols: 4 patients on the haplo-1 protocol had slowly falling chimerism and return of SCD, even as far as 10 years post-HCT; two patients on the haplo-PC protocol had falling chimerism with SCD recurrence. Another 2 patients have falling chimerism and may experience graft failure despite prolonged courses of immunosuppression. In the setting of mixed and waning chimerism some patients have continued immunosuppression for years despite the absence of cGVHD. Future studies to determine when tolerance has been achieved and immunosuppression can be safely discontinued are important.

Repeat HCT with myeloablative conditioning and stem cells from the original donor for those with falling but persistent DMC and return of SCD is being investigated with an ongoing study (NCT04008368). This strategy employs busulfan-based conditioning with the addition of serotherapy with aletuzumab, theorizing that immune tolerance was achieved in the first transplant, as evidenced by the persistent DMC, but adequate space must be created in the marrow for enduring engraftment.

The improved graft rejection rates associated with the PC regimen came at the cost of increased toxicity. Specifically, higher rates of viral reactivation and EBV-driven PTLD. A 2022 survey of international transplant centers regarding EBV infection demonstrated an overall frequency of EBV-PTLD of 1.28% in adults and 2.38% for haploidentical HCT with PT-Cy [48]. Despite the exclusion of EBV seropositive donors with seronegative recipients, the rate of EBV-PTLD was 23% on the haplo-PC protocol. Ultimately, EBV-PTLD contributed to death in 40% (2/5) of PTLD patients on the haplo-PC protocol. A similar pentostatin-cyclophosphamide pre-conditioning regimen with half the pentostatin dose was recently reported in the setting of matched sibling HCT for SCD [47]. In that setting, higher viral reactivation rates were reported with the addition of preconditioning without PTLD. In the era of regular PCR monitoring of EBV and pre-emptive treatment with rituximab, 30% mortality has been reported with PTLD [49].

This exceptionally high risk of PTLD and other complications as described above led to the early closure to enrollment on the haplo-PC protocol in 2023. Protocols employing PC may consider pre-transplant rituximab; evidence suggests a single dose given prior to HCT can significantly mitigate the risk of both EBV reactivation and PTLD with minimal toxicity[50].

Further, immune-mediated cytopenias were noted in four patients on the haplo-PC protocol, in two of whom it was refractory to treatment and resulted in death; no such complications were seen on the haplo-1 protocol. The pathogenesis of post-HCT cytopenias has not been completely elucidated, but delayed immune reconstitution post-HCT has been proposed[51]. Interestingly, our data do not demonstrate a difference between the protocols in total lymphocyte, CD4, CD8, or B-cell reconstitution post-HCT. Importantly, the addition of PC does not seem to impact the timing of hematologic reconstitution or increase the likelihood of febrile neutropenia, sepsis, or infection. Although viral reactivation trended up with the addition of PC, viral disease rates were unchanged, highlighting a potential implication of increased lymphodepletion. Patients with more refractory disease affecting multiple cell lines both had mixed lymphocyte chimerism at the time of diagnosis. Others have reported significantly lower CD3+ T lymphocyte donor chimerism at day +180 post-HCT in those who develop immune cytopenias [44]. We amended our protocol to increase myeloablation, changing from 400cGy given in 2 fractions to 400cGy given as a single fraction, with a goal of maintaining higher donor chimerism to help mitigate the risk of these immune-mediated complications and late graft failure. We hypothesized that the increased marrow suppression of the single fraction of TBI would give the donor cells an advantage and thereby improve and sustain donor chimerism, thereby mitigating immune complications seen more in mixed chimera patients[52]. Further adjustments may be considered in the future, including PC dose reduction. Others have employed two to three doses of pentostatin, with or without cyclophosphamide, with excellent engraftment rates and no reported PTLD or immune cytopenias [53-55]. While no patients on the haplo-PC protocol have developed a myeloid malignancy, enthusiasm should be dampened by the shorter period of median follow-up compared to the haplo-1 study. However, the lower graft rejection rate and higher incidence of full donor chimerism in haplo-PC compared to haplo-1 may have contributed to a lower risk for myeloid malignancies.

## Conclusion

Nonmyeloablative haploidentical HCT for SCD with PC preconditioning decreased acute graft rejection and long-term engraftment rates with minimal GVHD; there was also a trend towards improved DFS when compared to the haplo-1 regimen. While the original haploidentical trial had unacceptably high rates of graft rejection and myeloid malignancies that were mitigated in the haplo-PC protocol, complications associated with the haplo-PC protocol include viral reactivation, PTLD, and immunohematologic complications. The NIH enrolls patients with more severe disease compared to more standard haploidentical protocols, so results should be interpreted considering this. A novel nonmyeloablative haploidentical regimen for patients with SCD and compromised organ function is desperately needed.

## Acknowledgements

We would like to express our appreciation to Drs. Tom Hughes, Deb Citrin, David Stroncek, Bill Flegel, Luigi Notarangelo, as well as NP Donna Chauvet and PA Shawn Chong, for their support in implementing the protocol and providing clinical care to our patients.

## Authorship Contributions

EL, MH, NJ, and CF conceived and designed the analyses; EL, JV, JB, and WC collected data; JQ and MLG contributed data; EL and NJ performed the analyses; EL and CF drafted the manuscript. All authors reviewed and approved the final manuscript.

## Data Availability Statement

Data will be deposited in the Figshare data repository, DOI: 10.25444/nhlbi.29963303

---

**Table 1.** Baseline demographic and clinical characteristics SCD: sickle cell disease; TRV: tricuspid regurgitant velocity; ACS: acute chest syndrome; ICU: intensive care unit; RBC: red blood cell; HLA: human leukocyte antigen

**Table 2.** Transplant Outcomes at last follow-up GVHD: graft-vs-host disease; PTLD: post-transplant lymphoproliferative disorder; SCD: sickle cell disease; AIHA: autoimmune hemolytic anemia *one additional patient on the haplo-1 protocol developed myeloid malignancy post-HCT; this patient was excluded from formal analyses as she did not receive PT-Cy. Adjusted totals including those patients are as follows: haplo-1: 3/21 (14%); total: 3/42: (7%). ^one additional patients on the haplo-PC protocol developed PTLD; this patient was excluded from formal analyses as this was her second transplant. Adjusted totals including this patient are as follows: haplo-PC: 5/22 (23%); total: 5/40: (12.5%).

